# Knowledge + Innovation = Power: a protocol for implementing Aboriginal data sovereignty in an Aboriginal Medical Service for service delivery reform

**DOI:** 10.1101/2025.07.05.25330937

**Authors:** Emma Haynes, Lesley Nelson, Heather D’Antoine, Judith Katzenellenbogen, Elizabeth Armstrong, Lindey Andrews, Jasmin Brown, Nicole Bowser, Roz Walker, Ingrid Stacey, Jessika Purnomo, Dawn Bessarab

## Abstract

**Context:** Comprehensive Primary Health Care (CPHC) is an interconnected, holistic, and strengths-based health and wellbeing approach fundamental to Aboriginal Community Controlled Health Organisations (ACCHOs) in Australia. However, policy and funding trends increasingly threaten the capacity of ACCHOs to deliver CPHC by imposing burdensome administrative and accountability requirements. A central constraint is the marginalisation of culturally informed data measures and tools that could enable improved development, implementation, evaluation and reporting of CHPC services and programs. To mitigate this, ACCHOs must be enabled to take charge of collecting and using meaningful data to increase self-determination and drive impactful policy and service delivery. Central to this approach is the capacity, skills, and infrastructure to collect and use culturally informed data and tools that align with community needs and organisational imperatives.

**Materials and Methods:** This project will undertake an Aboriginal participatory action research (APAR) mixed methods developmental evaluation approach to Aboriginal Data Sovereignty (ADS) initiatives. The project will be conducted both centrally and across the regional sites of the South West Aboriginal Medical Service (SWAMS), an ACCHO in regional Western Australia. Activities to initiate the APAR process include the development and training of a Community Research Panel to lead/facilitate APAR projects. A series of regional workshops will be held to build data literacy (including regional population health data) and identify innovative culturally-informed health and wellbeing data measures and tools for selected health priorities. Project team Aboriginal academic researchers and Aboriginal researchers at SWAMS will ensure the inclusion of Aboriginal research methods (Aboriginal ways of knowing, being and doing). Data collection will include both quantitative and qualitative data which will be analysed to identify the enabling processes and community and organisational outcomes as they align to the principles of ADS.

**Discussion and next steps:** The protocol herein describes the first phase of a two-phase project, where the second phase will implement the new and/or adapted data measures and tools established in phase one of the project. This project will build capacity towards evidence-based decision making by ACCHOs and support self-determination by enabling local, real-time evaluation of the integrated models of care that ACCHOs deliver.

## Introduction

### Context

*“In the absence of meaningful data, Indigenous people the world over are creating their own metrics. Built on the foundations of Indigenous cultures, languages and science”* (Mokak, 2024).

#### Terminology and Language Use

**Aboriginal** is used throughout this paper to refer to the First Peoples of Western Australia. This aligns with community preferences and Western Australian Government data standards:

*“The term Aboriginal people is used in preference to ‘Indigenous’ or ‘Aboriginal and Torres Strait Islander’ people, in recognition that Aboriginal peoples are the original inhabitants of Western Australia.”*

When referring to populations across Australia, we use **First Nations** to acknowledge the sovereignty and diversity of Aboriginal and Torres Strait Islander peoples nationally.

We use **Aboriginal Data Sovereignty** in line with WA policy and local usage. While **Indigenous Data Sovereignty** is nationally recognised (e.g., by the Maiam nayri Wingara Collective), we adopt “Aboriginal” to reflect the terminology used in our context.

When citing others, we retain the original terminology used in their work to respect the context and intent of the source.

In the Australian healthcare context, Comprehensive Primary Health Care (CPHC) is an integrated and culturally sensitive approach encompassing delivery of a wide range of services across the physical health, social and emotional wellbeing, prevention-treatment continuum, including: clinical, disability, allied health, child and maternal health, social and emotional wellbeing, mental health, alcohol and other drugs, dental services, and community programs (1). The CPHC focus on community involvement and recognition of historical, economic, social, and cultural determinants of health is a holistic approach to primary healthcare highly valued by Aboriginal clients. It is important for Aboriginal community-controlled health organisations (ACCHOs) to have the authority, autonomy and resources to be able to put community ideas into action, building on their knowledge and innovation to increase capacity for CPHC service delivery (2, 3).

However, short-term funding models often do not support the CPHC focus on long-term and continuous work with the community to achieve its goals, outcomes and evaluation (4). That is, current government policies, funding mechanisms and associated reporting and performance indicators often fail to support or reflect the priorities of delivering CPHC, creating tensions between the community need for holistic and culturally sensitive healthcare and government policies and funding mechanisms that determine the delivery models (4, 5). Three main themes that contribute to these tensions include: 1. an emphasis on quantifiable metrics that overlook the holistic and culturally sensitive nature of the care offered;2. prioritising standardised outcome measures not aligned with the wellbeing outcomes valued by Aboriginal communities; and 3. reporting to meet funders’ requirements that diverts attention away from meaningful planning, implementation, and evaluation of holistic services. Further impacting on ACCHOs ability to deliver CPHC services is the burden of reporting when services are delivered using multiple separate funding streams, from smaller grants to longer-term funding contracts (6).

Funding pressures negatively impact health promotion and social and emotional wellbeing (SEWB) activities; with consequences including short-term, limited programs, challenges to the creation of a stable, qualified health promotion workforce, and a lack of program evaluation crucial for evidence-based decision-making, program improvement, and continuation of government funding (7). This funding short-fall and workforce insecurity is significant in its lack of alignment with CPHC principles where a focus on cultural, holistic health and empowerment of community through self-determination is fundamental to SEWB.

The challenge here is that mainstream population health indicators used to measure the health and wellbeing of First Nations people are constrained by the Western biomedical paradigm, which focuses mainly on deficit and disease and understands wellbeing primarily at an individual level. In contrast, First Nations peoples’ views of wellbeing encompass more complex, interconnected, holistic, relational and strengths-based concepts of wellbeing at multiple levels (8). Thus, there is often limited data/evidence to illustrate what works (or does not work), for who and why, with most Aboriginal-specific programs not evaluated to assess whether they achieved their goals and objectives and fewer still using culturally appropriate methods for assessing program effectiveness. This lack of culturally responsive evidence is the result of the dichotomy in the commissioning of Aboriginal health and wellbeing program evaluations between “what the Funder wants versus what the Community needs” (9). To advance First Nations self-determination within the healthcare system, population health reporting must be transformed to reflect First Nations peoples’ ways of knowing and views of health - incorporating perspectives, values, and cultural contexts to inform how health data is collected, analysed, and used. This transformation requires greater focus on cultural safety, cultural responsiveness, and collaboration with First Nations communities.

The ACCHO sector increasingly recognises the importance of mitigating the impact of sustained financial pressure, regulatory constriction on health services and outcomes, and the risk of tending towards a more neoliberal mode of operation emphasising market-driven approaches that minimises government spending. ACCHOs must develop and implement innovative, adaptive strategies (10), including collecting and using meaningful data in a way that increases self-determination, wellbeing and drives impactful policy and service delivery (11). While there has been much consultation and research to identify community aspirations and desires for service delivery, the knowledge translation challenge is to implement learnings into practice. This reflects the need within health services for innovative culturally-informed data measures and tools that align with both community health priorities and organisational imperatives (12, 13). Building meaningful data into the operation of administrative systems (data ***for*** governance) and concurrently ensuring data governance structures, procedures, and processes are developed by First Nation peoples (governance ***of*** data) has the potential for profoundly positive impacts.

It is essential that First Nations peoples have ownership over their data to determine their own future and ensure data reflects their realities and priorities (14). Indigenous data sovereignty (IDS) is an international movement, that embodies the sovereign right of Indigenous peoples to regulate, collect, use, and own the data that is recorded about their people, communities, resources, and lands (14, 15). The IDS movement has arisen to ensure the rights to self-determination through access to and use of data to advocate for, improve and maintain Indigenous peoples’ overall well-being (14). In Australia, the Maiam nayri Wingara Indigenous Data Sovereignty Collective and the Australian Indigenous Governance Institute collaborated to articulate five key IDS principles (16) that have become widely accepted as foundational criteria in assessing IDS readiness and capacity (17). These principles represent the right for First Nations communities to decide which data require active governance and to participate in data processes that align with these principles:

1. Exercise control of the data ecosystem including creation, development, stewardship, analysis, dissemination and infrastructure.
2. Data that are contextual and disaggregated (available and accessible at individual, community and First Nations levels).
3. Data that are relevant and empowers sustainable self-determination and effective self-governance.
4. Data structures (eg protection and use) that are accountable to Indigenous peoples and First Nations.
5. Data that is protective, strengths-based and respects Indigenous interests, including after dissemination.

The principles of IDS (hereafter Aboriginal data sovereignty (ADS)) have been embedded in practical resources such as the Lowitja’s IDS evaluation toolkit (18) and in First Nations health policy through the National Agreement on Closing the Gap (NA-CtG) Priority Reforms (19, 20) with corresponding state-based action plans and evaluation frameworks. These policies emphasise giving First Nations people a genuine say in the delivery of services through shared decision-making on the design, implementation, monitoring and evaluation of policies and programs (19). The Priority Reforms are intended to drive structural change to the way decisions are made, and funding is allocated. They also commit governments to strengthen community-controlled organisations as they “deliver healthcare services that are holistic, comprehensive, and culturally competent and better for our people. They get better outcomes, and they employ more Aboriginal and Torres Strait Islander people” (19, 20). Thus, the NA-CtG includes an agreement by government bodies to identify opportunities for reprioritisation of funding towards Aboriginal community-controlled organisations. Of particular relevance to ADS, NA-CtG Priority Reform 4 aims to ensure that First Nations people have access to, and the ability to use, locally-relevant data and information to drive efforts to reform healthcare (21). The application of NA-CtG reforms to the ACCHO sector may be particularly useful in mitigating detrimental policy and funding trends and ensuring implementation of research findings reflecting community aspirations and desires, thereby shifting the power balance in research.

Responding to the expressed needs of communities to enact ADS in practise, we have developed a two-phase project to explore a mechanism for strengthening CPHC in the ACCHO sector (see Fig 2). This protocol paper describes Phase 1 of the project that aims to develop and evaluate capacity, skills, and infrastructure for collecting and using culturally informed data and tools aligned with Aboriginal community needs and organisational imperatives. There are three Phase 1 project objectives:

1. Build and evaluate confidence, skills and knowledge about access to and use of population health and wellbeing (HWB) data;
2. Review, co-develop and evaluate innovative culturally-informed HWB data measures and tools (both clinical and non-clinical) for selected health priorities;
3. Report on operational enablers and barriers for implementation of new data measures and tools for selected HWB priorities for the Phase 2 project.

This project will thus provide a case study demonstrating how the HWB priorities of Aboriginal communities can inform measures and indicators that can be applied by services to their models of care and operational management. The project is based in the South West Aboriginal Medical Service (SWAMS) an ACCHO located in the South West region of Western Australia. The study will be set in and around the SWAMS region, including eight communities where SWAMS has clinical sites. This covers an area of around 40,000kms^2^ (See Fig 3) and includes the following clan groups: Pinjarup; Wardandi; Wilman; Kaniyang; Bibbulman and Minang.

## Materials and Methods

### Position statement

The formal partnership developed for this project between the University of Western Australia (UWA), SWAMS and Wungening (a large Perth-based Aboriginal Community Controlled service provider) consolidates many years of experience in combining data literacy, critical thinking and Aboriginal control of research/data with improved community HWB outcomes. The strong academic track record of the team in developing and facilitating Aboriginal community-determined action research projects and use of data is combined with the extensive experience of SWAMS and Wungening in culturally-led service delivery and community engagement.

### Theoretical and conceptual framework

We propose to undertake an Aboriginal participatory action research (APAR) mixed methods developmental evaluation of activities undertaken to build ADS (Fig 1). Our methodological approach is informed by standpoint theory which acknowledges and prioritises the perspectives of First Nations peoples to create more objective accounts of the world from their position (22, 23), and decolonising theory which aims to reduce the detrimental effects caused by colonisation (24, 25). This theoretical lens informs our research methods that are based in Aboriginal ways of knowing, being and doing including both way learning, building community research capacity, co-design with communities, and holistic health approaches (26–28).

**Fig 1.** Aboriginal Participatory Action Research (APAR) cycle beginning with identification of priorities.

APAR is a strengths-based approach focusing on activating existing power, developing abilities and critical thinking skills to facilitate action for change, in turn contributing to a greater sense of control and wellbeing (29). APAR is particularly relevant where health disparities are the result of marginalisation, colonialism and power differences by facilitating communities to collectively gain greater influence, agency and control over their health. APAR is an ideal fit to achieving aspirations of ADS at SWAMS.

The empowering potential of APAR, building community research capacity through participation in a skilled and resourced research advisory group, provides a Theory of Change framework for our project (an outline of how and why the changes are expected to happen), guides evaluation questions and promotes a more nuanced analysis of findings (exploring the how and why, or why not) (30–32).

### Project Aim

The project will develop and evaluate actions intended to deliver the capacity, skills and infrastructure required for collecting and using culturally-informed data and tools that align with community needs and organisational imperatives.

### Overall study design

We will combine qualitative insights that capture lived experiences and perceptions with quantitative measures. Such mixed methods approaches provide a more holistic and nuanced interpretation of the complexities involved and ensure the inclusion of a broad scope of information, allowing for triangulation of data, particularly when process indicators are linked to an appropriate theory of change.

A series of activities will contribute to the Phase 1 objectives, each being evaluated and informing the development of the data measures and tools.

### Actions (Implementation)

A Community Research Panel (CRP) will be established to provide opportunities and a structure for community members to lead the APAR process. The recruitment of the CRP members will be through regional community consultations and an Expression of Interest process aiming for diversity of experiences, opinions, gender, age, and family groups. The project team will consist of Aboriginal academic researchers and Aboriginal researchers at SWAMS who will facilitate the Aboriginal research methods (ways of knowing, being and doing) component of the CRP workshops. The CRP will be convened and facilitated by an Aboriginal researcher employed at SWAMS as the project manager. This will allow for ongoing skill development throughout the CRP meetings, in addition to the workshops. The CRP members will also be funded to attend additional capacity building opportunities, such short courses provided by the Lowitja Institute (28) (Australia’s only Aboriginal and Torres Strait Islander community-controlled health research institute).

The CRP will also work closely with the SWAMS Research Advisory Group, an existing body established to guide the governance of research activities conducted at SWAMS, its membership is primarily SWAMS staff plus two external researchers (See Fig 4). In collaboration with the Research Advisory Group, the CRP will provide input to help assess research applications and to determine research priorities based on knowledge of community needs, service gaps and health priorities. This will allow SWAMS to make more informed decisions when choosing to accept or decline external requests to conduct research involving SWAMS. The CRP will also support the need for community leadership in delivery of SWAMS programs and services.

A series of activities will contribute to the Phase 1 objectives, each being evaluated and informing the development of the data measures and tools. Central will be the design and delivery of three workshops, conducted both centrally and across SWAMS 8 sites: 1) ADS data literacy workshops; 2) Top five health priorities workshop; and the 3) Measures for implementation workshop. These are described in more detail below.

#### 1) ADS data literacy

Initial “Demystifying data’’ workshops across the regional areas where SWAMS operates will include activities and discussion aimed at increasing participants skills, knowledge, confidence and capacity in understanding and working with data. Trust in data will be strengthened through improved understanding of its sources, interpretation, and application. We will seek the assistance of data custodians for the extraction of descriptive quantitative health-related data pertaining to the SWAMS catchment area from a range of routinely available population-based administrative data sources. Subsequently, mechanisms will be co-designed with the CRP for sustainability and accessibility of data sharing with and by government agencies.

For the workshop, population health data will be collated and presented in accessible formats, with discussion of its use in evaluation, service development, policy and advocacy. Presenting current population health data will highlight health priority areas determined by the government. Workshop activities will facilitate discussion amongst CRP members to share their opinions on whether government priority areas align with community perceptions of priority areas. Learnings from the Wungening Aboriginal Corporation experiences will provide practical current examples (33) including in the use of data in advocating for policy and funding model changes. Additional topics include the importance of ensuring Data Governance and Stewardship, exploring the concepts around data protection, integrity, responsible use, and accountability. The workshops will also foster a greater understanding of Aboriginal research methodologies, including Aboriginal ways of knowing, being, and doing.

#### 2) Determining health priorities

Guided by the CRP and sampling for diversity, we will engage key stakeholders (community leaders, healthcare providers, and members) to undertake surveys regarding community-perceived health needs and priorities across the SWAMS region. Following a summative analysis of the surveys, the top five health priorities for the project will be determined through a consensus workshop with the CRP, the project team, and the SWAMS Board and staff members.

#### 3) Determining measures for implementation

Within the selected health priority areas the project will review, co-develop and evaluate innovative culturally-informed HWB data measures and tools (both clinical and non-clinical) through two mechanisms. First existing SWAMS data collection tools and data sources will be investigated for relevance to the five selected health priorities and possible new ways to analyse and utilise this existing data will be explored. Secondly, we will consider the development of new, innovative data sources and tools.

Additional project activities include a narrative literature review to identify the key facilitators and barriers to implementation of community-driven health and wellbeing measures have been implemented in Aboriginal community-controlled health services. This includes examining how community-identified health and wellbeing priority areas are identified and translated into practice. The review will also examine the relationships and distinctions between community-driven measures and funding body KPIs. This involves comparing how community-driven measures and CQI processes as opposed to externally imposed outcome measures influence service design, delivery and uptake, and the extent to which they align- or conflict-with community preferences.

Finally, selected SWAMS staff members supported to undertake a “deep dive” regarding the development of outcome measures in topics related to the identified priority health areas. as with the literature review this exploration will contribute to the development of possible new ways to analyse and utilise existing data, as well as potential new and innovative data sources and tools.

### Data collection

All project participants will contribute to the iterative, developmental evaluation. Following standard consent processes, the sample is anticipated to include CRP members (n = ∼12), community workshop participants (n = ∼30), other community members within Bunbury and the SWAMS regional areas (n = ∼ 30), SWAMS staff (n = ∼10), CEO and Board members, and project team members (n = ∼12). The CRP and SWAMS Aboriginal project manager will guide creation of survey questions, and interview prompts and data analysis for the developmental evaluation of the processes/implementation and outcomes of activities. CRP members will have access to project team Aboriginal academic researchers and SWAMS Aboriginal research staff to advise regarding Indigenous data collection methods. Below are the anticipated data collection tools though these may be modified by the CRP (see Appendix.

1. A pre- and post-survey of regional community workshop participants and CRP members, using Likert scales and open-ended responses to questions regarding research knowledge and confidence (e.g. regarding data collection, analysis, reporting and advocacy), perception of local HWB priorities (needs and aspirations), knowledge of population health data and data custodians, and the value and accessibility of population health data visualisation resources.
2. Yarns/yarning circles (interviews/focus groups) (34) with CRP members to explore their experiences, issues in implementing the CRP, most important outcomes, learnings, and perceptions of impact on SWAMS policy and practices. We will also conduct yarns/yarning circles with SWAMS staff, including managers and board members to assess to what extent CRP decisions impact SWAMS policy and practices, and to identify enablers and barriers to implementing CRP decisions and priorities including systemic issues such as data tools and existing measures. Surveys, yarns and yarning circles will be utilised to evaluate the implementation of workshops and the outcomes they achieved.
3. The Project Journal, which will be the shared responsibility of all project staff. The journal will record unintended consequences and issues in implementing the CRP, the most important outcomes, learnings, perception of impact on SWAMS policy and practices. The journal will also capture observations regarding the evolving nature of the project and issues related to the day-to-day planning and implementation of the project not otherwise captured by meeting minutes.

As all individuals directly involved in the implementation of the project will be invited to participate, the sample is purposive and fixed, rather than driven by theoretical sampling. In this context, data saturation does not need to be determined. Participant recruitment will commence in the second year of the project (December 2025) and data collection will take place over a period of nine months (completing July 2026). All surveys, yarns and yarning will be conducted in English, electronically recorded (unless requested to be taken as notes only) by nominated co-researchers (See Appendix 1. Preliminary survey and Yarning questions for more detail). Informed consent will be provided in writing.

Data will be secured and safely stored according to the University of Western Australia policies and protocols. In accordance with participant consent, de-identified or anonymized data underlying reported findings will be deposited in an appropriate public data repository such as the Qualitative Data Repository Data (https://qdr.syr.edu/). Noting that while not directly identified in some cases data will be collected from a small group of participants and may not be shared if it contains indirect identifiers (such as sex, ethnicity, location, etc.) that may risk the identification of study participants.

### Data Analysis

Data analysis and write up will be iterative and guided by the CRP, ensuring results support collective interests. The findings regarding the enabling processes at community and organisational levels will be analysed for alignment with the principles of ADS. Quantitative and qualitative analysis will be used to assess various outcome measures such as level of staff research skills, knowledge and capacity, level of understanding and trust in data and continued relationships with data custodians. Analysis will also involve description, synthesis, and critical review of the extent to which reviewed measures were feasible, useful and implemented. Additional translational outcome measures will include total number of outcomes achieved, such as models of care developed, signed research agreements between SWAMS and researchers, research staff employed, enrolments and completions of research-related short courses, recommendations translated into policies, and lay and academic publications.

Learnings will be synthesised and presented as an Implementation Report regarding operational challenges and solutions regarding potential innovative measures in the prioritised health domains. Report recommendations for Phase 2 implementation will cover enabling processes such as capacity building, organisational systems and structures, and data infrastructure as well as technical solutions required for optimal use of data for ADS. The recommendations will also be reviewed for the extent to which the new measures/ processes contribute to SWAMS organisational imperatives and aspirations. That is: 1) to develop and implement sustainable and culturally-safe model of care, care coordination and operational management 2) to use SWAMS own data to advocate for funding and policy; and 3) to build organisational and local research capacity. Project write up will highlight key impacts from a proof-of-concept project, including consideration of the ethical implications. We will also publish the findings as a peer-reviewed journal article.

### Ethics

Ethics approval has been received from the Western Australian Aboriginal Health Ethics Committee (HREC1407). As an ADS project strong processes and protocols will be established at SWAMS to guard against unethical collection and use of data. A comprehensive governance and reporting structure has been established to support the project reflecting the aspirations of ADS and Aboriginal leadership (see Fig 4) and is consistent with NHMRC ‘Keeping Research on Track’ requirements (35). This includes appropriate participant recruitment and consent processes.

### Findings - developing Phase 2

The project will take place between 2025-2026 and will form a foundation for Phase 2 of the project, where the new health and wellbeing data measures and tools will be implemented, as described in Fig 2.

**Fig 2.** Relationship between Phase 1 Aboriginal Participatory Action Research (APAR) cycle beginning with identification of priorities and Phase 2 APAR cycle.

## Discussion

Insights from our project are anticipated to include the enablers and barriers to implementing new data measures and tools, types of health and wellbeing measures that are important to First Nations peoples. Project benefits include capacity and confidence building for ACCHO staff and community members around research and ADS principles.

In addition to supporting Phase 2, our findings have the potential to act as a guide for the ACCHO sector, funding bodies and data custodians, emphasising the value of ADS, as well as key insights and nuances when incorporating ADS within an organisation. The NA-CtG requires structural change, giving First Nations peoples a genuine say in the delivery of services encompassing shared decision-making on the design, implementation, monitoring and evaluation of policies and programs (20). The project aligns with NA-CtG. Priority Reform areas 2. Building the Community-Controlled Sector and 4. Shared access to data an information at a regional level. Our project is well placed to impact NA-CtG priority actions such as shared decision-making with governments to accelerate policy and place-based progress (19).

Through alignment with the NA-CtG we see this project contributing to reforming the systemic and structural racism often imposed on ACCHOs. Capacity building opportunities (such as operationalising ADS, mobilising data literacy, and developing the healthcare research skills, capabilities and leadership of the CRP, wider community, and staff) will allow ACHHOs to prioritise and value community participation, a culturally appropriate and skilled workforce and self-determination (4). This in turn will improve the accessibility and cultural appropriateness and responsiveness of services, as well building trust within the community (36).

## Conclusion

This project will address a knowledge gap regarding the enablers for implementing culturally appropriate measures into service design and models of care, and addresses challenges such as achieving a balance between measures tailored to reporting and accountability and those reflecting community priorities (37). This Aboriginal-led, capacity building action research project addresses an area of unmet need for innovative culturally-informed data measures and tools that align with both community health priorities and organisational imperatives. The Aboriginal community-controlled sector increasingly recognises the importance of collecting and using meaningful data in a way that increases self-determination, wellbeing and drives impactful policy and service delivery. However, gaps persist in achieving this vision due to challenges including: emphasis on quantifiable, non-holistic metrics; prioritising standardised outcome measures; and excessive reporting to meet funders’ requirements.

The recommendations from this research will be directed at funding bodies and policy makers for consideration when designing funding models and administrative and accountability requirements for ACCHOs, including taking into consideration the differences in the types of HWB measures and tools that are required to meet the needs and priorities of Aboriginal peoples. Additionally, the results of this project could be used as proof-of-concept for implementation of ADS into the organisation and service delivery models of ACCHOs nationally.

## Data Availability Statement

As this is a study protocol, datasets have yet to be generated or analysed. This article does not report data. Given the nature of certain confidential and/or identifiable data that will be collected, some data will have restricted access. Data will be de-identified and made available following study completion. We plan to disseminate data and study findings through peer-reviewed publications, community reports, and a report to SWAMS regarding implementation of Phase 2.

## Supporting information

**S1 Fig 1.**
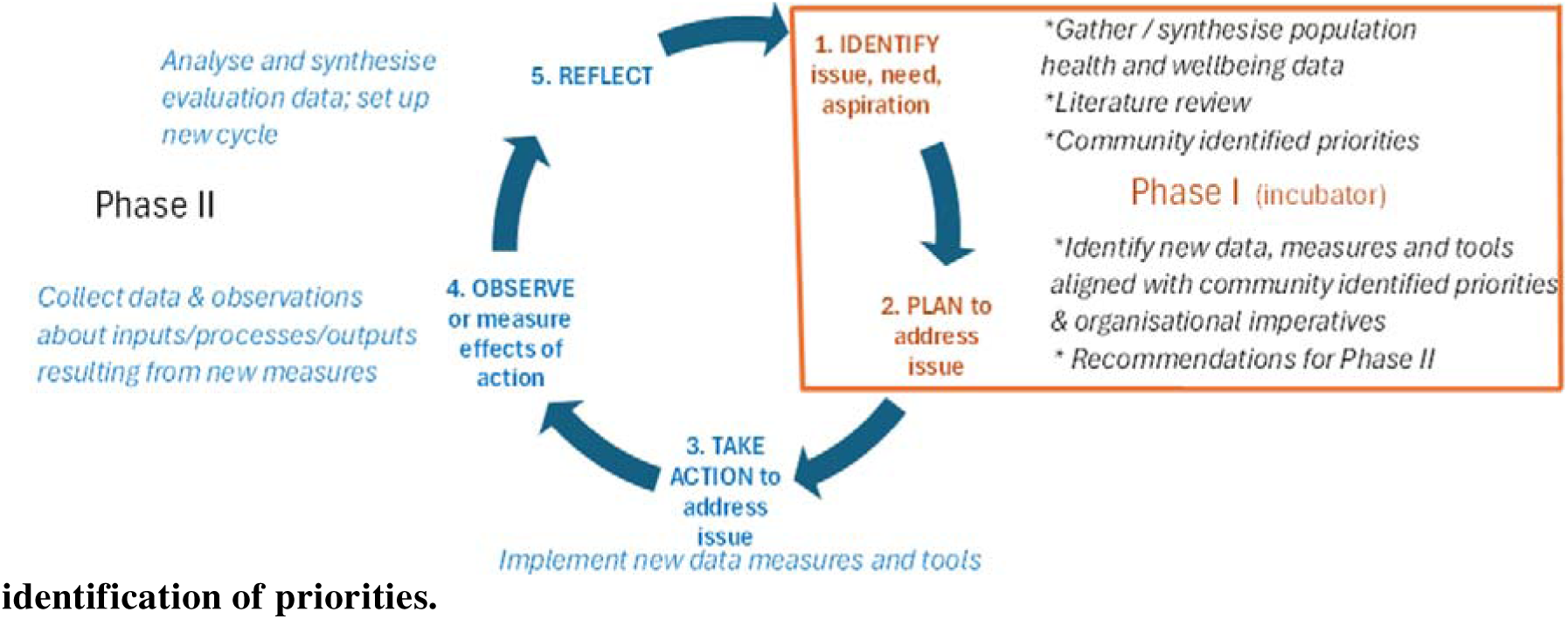
Aboriginal Participatory Action Research (APAR) cycle beginning with identification of priorities.

**S2 Fig 2.**
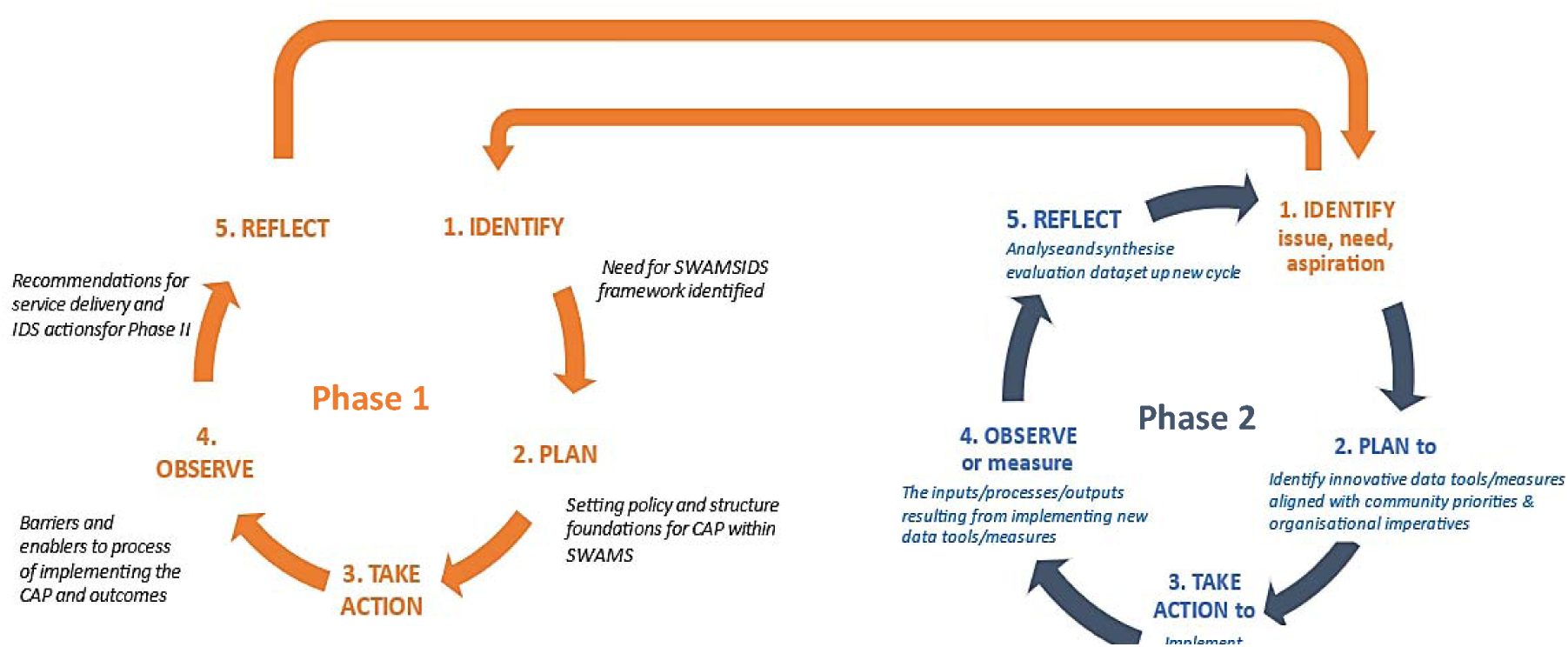
Relationship between Phase 1 Aboriginal Participatory Action Research (APAR) cycle beginning with identification of priorities and Phase 2 APAR cycle.

**S3 Fig 3.**
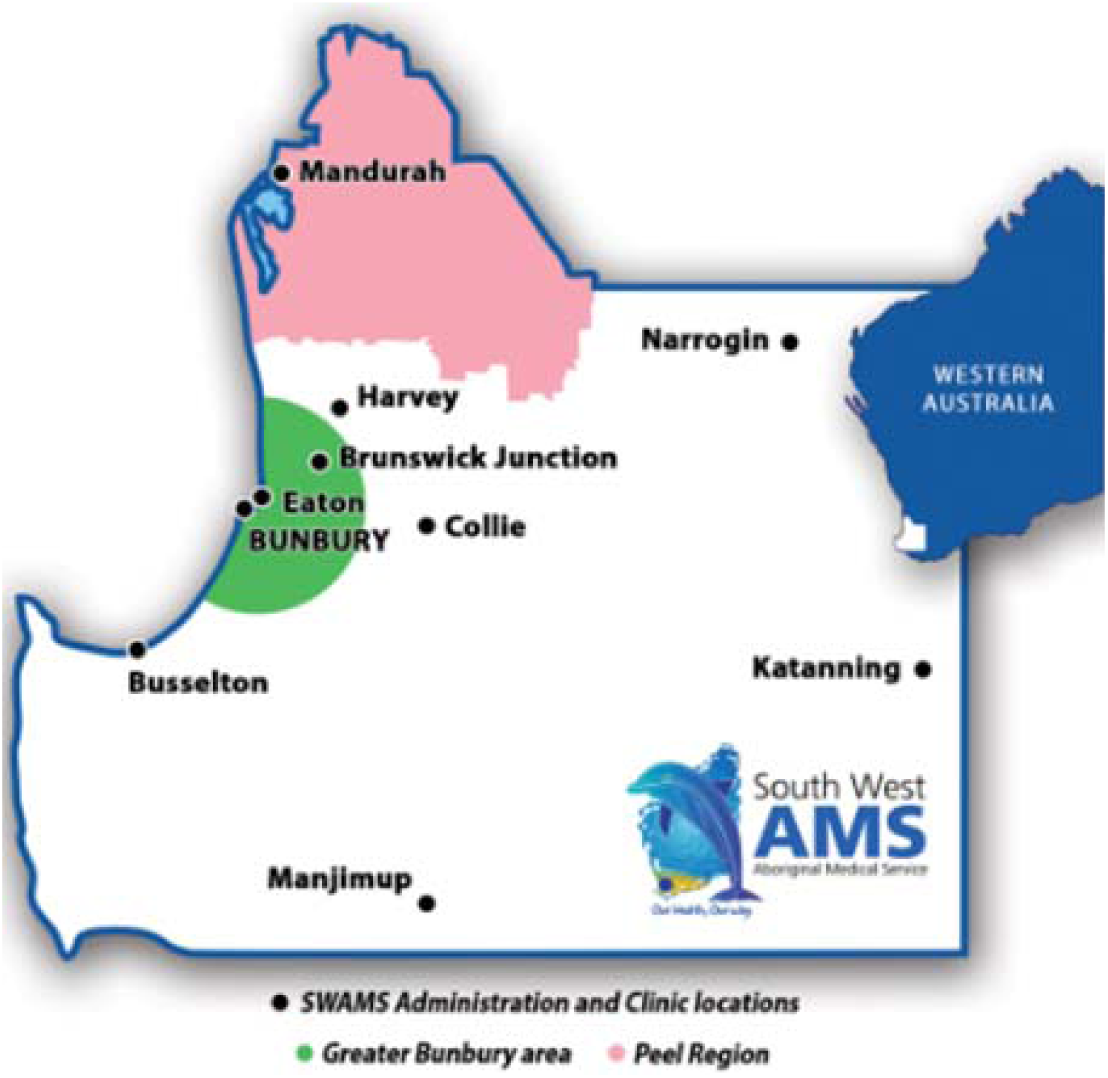
Map of SWAMS clinical sites.

**S4 Fig 4.**
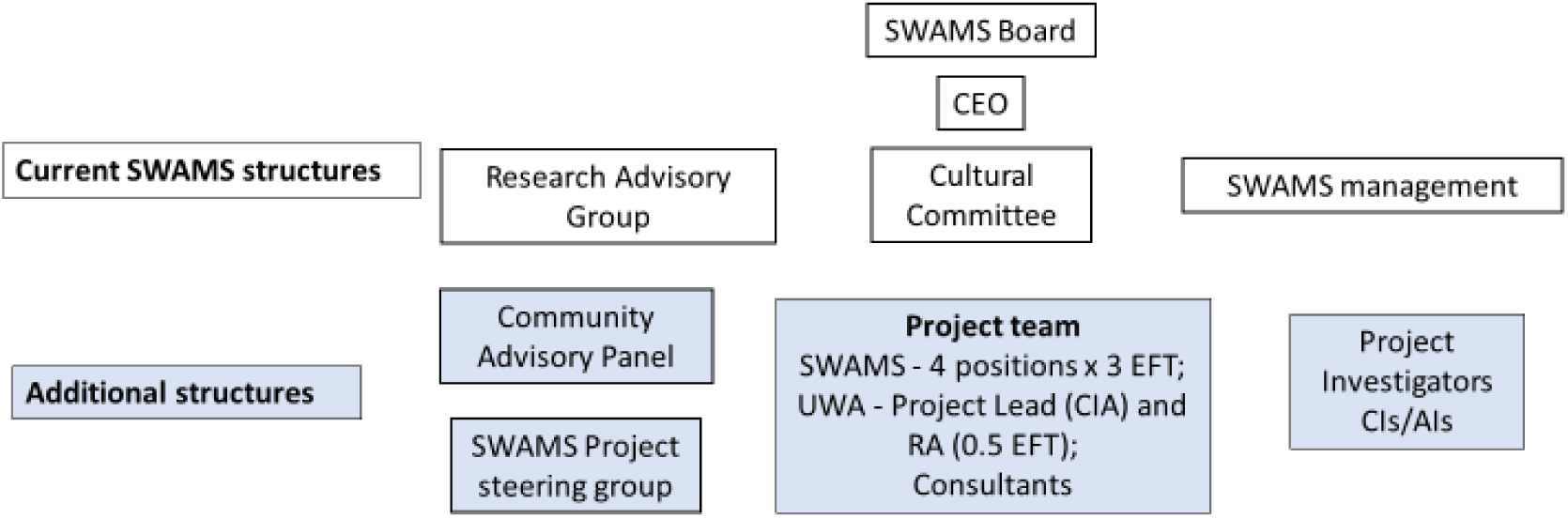
Governance and reporting structure within SWAMS.

## Appendix 1. Preliminary survey and Interview questions

(Final questions to be determined by the Community Panel co-researchers and SWAMS Research team)

### Survey questions

- Likert scale questions

o To what extent has your knowledge of research increased?
o To what extent confidence in discussing research increased?
o To what extent has your knowledge of data collection, analysis, reporting and advocacy increased?
o To what extent has your understanding of local HWB priorities/needs increased?
o To what extent has your understanding of local HWB aspirations/hopes increased?
o To what extent has your knowledge of population health data and data custodians increased?
o Did you feel that the population health data resources provided were easy to understand?
o Did you feel that the data visualisation resources provided were useful?
- Open-ended responses – free text spaces below each of above questions plus an additional space for “Other information you would like to tell us”

### Interview and yarning circle questions

#### 1. Community Research Panel

Preliminary yarn regarding extent of involvement.

- Can you tell me/us about your experiences during the formative phase of the CRP?
- Can you tell me/us about your experiences during the process of setting up the evaluation research (determining research questions, how to conduct surveys etc)?
- Where there any challenges? Can you tell me/us more about those?
- What have been the most important outcomes/learnings for you? Can you tell me/us more about those?
- To what extent do you feel our work has had impact on SWAMS policy and practices? Or has the potential to? Can you tell me/us more about that?

#### 2. SWAMS staff, including managers and board members

Preliminary yarn regarding extent of involvement.

- Can you tell me/us about your perceptions regarding this project, including the formation of the CRP and its implementation of research?
- What do you see as the benefits/opportunities from having a CRP and its outcomes?
- Where there any challenges? Can you tell me/us more about those?
- What have been the most important outcomes/learnings for you regarding the CRP and its outcomes? Can you tell me/us more about those?
- To what extent do you feel the work of the CRP has had impact on SWAMS policy and practices? Or has the potential to? Can you tell me/us more about that?
- What do you see as the enablers and barriers to implementing CRP decisions and priorities including systemic issues such as data tools and existing measures?

